# Multilingual video education for hospitalised patients with Myocardial Infarction (EDUCATE-MI): single-arm implementation study

**DOI:** 10.1101/2025.07.01.25330381

**Authors:** Aileen Zeng, Edel O’Hagan, Sul Ki Kim, Simone Marshner, Mitchell Sarkies, Marina Bahgat Wassif, Christine Ji, Julie Ayre, Daniel McIntyre, Clara K Chow, Aravinda Thiagalingam, Liliana Laranjo

## Abstract

**Background:** International guidelines recommend that after a myocardial infarction (MI), patients receive education on key secondary prevention medications and lifestyle modifications, before hospital discharge. Multilingual videos may help deliver high-quality patient information to patients with diverse preferred languages and literacy levels, but little is known about the feasibility and impact on knowledge of this approach in the inpatient setting. To evaluate whether an educational video (available in English, Arabic, Hindi and Mandarin) provided to hospitalised patients post-MI can feasibly improve patient knowledge of MI.

**Methods:** This was an effectiveness-implementation hybrid quasi-experimental study. Recruitment took place between December 2023 to October 2024 in a tertiary hospital in Sydney, Australia. The intervention consisted of a 5-minute educational video on post-MI management, available in English, Arabic, Hindi and Mandarin. The primary outcome was change in patient knowledge of MI measured by comparing the mean number of correct responses before and immediately after the intervention using a two-sample t-test. Implementation was assessed by evaluating the acceptability and fidelity of the video. We performed thematic analysis on the notes taken of participants’ feedback for improving the video.

**Results:** Of the 129 participants recruited, 20.2% (26/129) were female and the mean age was 59.4 years (Standard deviation [SD] 12.6). For 74.4% (96/129) of participants English was their preferred language, with Hindi the predominant preferred non-English language (13.2%, 17/129). The average number of correct responses out of 10 at baseline was 5.4 (SD 2.7) compared to 7.2 (SD 2.5) post-intervention (mean difference = 1.9; 95% CI 1.6, 2.2, p<0.001). The educational video was well-accepted, with 83.6% (107/128) of participants finding it easy to understand, 74.2% (95/128) finding it engaging, and 87.5% (112/128) considering it useful. Fidelity of the intervention was assessed as high, as core components (i.e. animations and educational content via audio and subtitles) were delivered as intended. Themes from participants’ feedback for improvement include content complexity and preference for conversational language and dialects.

**Conclusion:** A MI educational video delivered during inpatient hospital admission, available in multiple languages, may improve patient knowledge in the short term. Further scaled research is needed to evaluate the effectiveness and implementation of this intervention in other hospitals and settings, and to assess additional languages and strategies to support the health education needs of diverse populations.

## Introduction

Acute myocardial infarction (MI) secondary to plaque rupture (type 1 MI) is a major cause of cardiovascular mortality, accounting for 38-44% of all cardiovascular disease (CVD) related deaths worldwide.^1^ After the acute phase, the risk of death from another cardiac event remains high.^2^ International guidelines recommend that patients receive education on key secondary prevention medications and lifestyle modifications prior to hospital discharge, with early initiation of these strategies following the index MI to reduce the risk of morbidity and mortality.^3,4^

Secondary prevention of MI involves cardiac rehabilitation, a combination of lifestyle modifications and adherence to different medications, including long-term aspirin, dual antiplatelet therapy (for 6 to 12 months), lipid-lowering medications, and beta-blockers.^5,6^ However, medication adherence remains suboptimal, with only 66% of patients adhering to all classes of drugs—such as aspirin, angiotensin-converting enzyme inhibitors, angiotensin receptor blockers, beta-blockers, calcium channel blockers, diuretics, or statins—used for secondary prevention at 12 months post-MI.^7^ Medication adherence is influenced by a combination of psychological factors, the complexity of the drug regimen and instances where prescribers fail to consider patient needs.^8^ Patient education is an important component in addressing suboptimal medication adherence.^9^

Educational videos can help deliver evidence-based health information to diverse populations and accommodate different language preferences and literacy levels, including people with limited reading ability or illiterate.^10,11^ A systematic review (59 experimental studies, n=9789) of video-based educational interventions reported improved knowledge in 75% (30/40) of the assessed outcomes in people with chronic disease.^10^ Previous studies evaluating inpatient video interventions for patients post-MI^12,13^ have shown improvements in patient knowledge, but the videos in these studies were delivered in just one language. At present, no studies have evaluated the impact of multilingual educational videos delivered during admission to improve patient knowledge post-MI.

The aim of this study was to assess the impact on patient knowledge and the feasibility of a multilingual educational video provided to hospitalised patients post-MI.

## Methods

### Study design

We conducted a type 1 effectiveness-implementation hybrid trial using a single-arm quasi-experimental design in a tertiary teaching hospital in Sydney, Australia. The protocol is available on <u>Open Science Framework</u> (registered on 12 April 2024). Reporting of this study follows the Standards for Reporting Implementation Studies (StaRI), and Transparent Reporting of Evaluations with Non-randomized Designs (TREND) reporting guidelines and Template for Intervention Description and Replication (TIDieR) checklist^14–16^ (Supplement 1-3).

### Ethical statement

Ethics approval (2021_ETH00983_v3) was obtained from Western Sydney Local Health District Human Research Ethics Committee. Participant information and consent forms were available in English, Arabic, Hindi and Mandarin. Informed consent was obtained in the participants’ preferred language. All documents and content in the intervention were translated by the multi-cultural unit in Western Sydney Local Health District by a qualified health interpreter.

### Participants and setting

Patients aged 18 years old or older with documented MI admitted for inpatient services at Westmead Hospital for a type 1 MI^17^ who had undergone coronary angiography and understood one of the four available languages: English, Arabic, Hindi and Mandarin were invited to participate (Supplement 4).

Participants who were unable to consent to the study in one of these four languages or were unable to complete the video due to a cognitive or sight impairment were excluded.

### Intervention and implementation strategy

The intervention was a single video, approximately 5 minutes long, with voiceovers and subtitles in English, Hindi, Arabic and Mandarin (subtitles in Simplified Chinese). The video included different animations and focused on explaining the disease process leading to a type 1 MI (i.e. acute coronary atherothrombosis) and the importance of different medications post-MI. The video was developed by a consultant cardiologist and cardiology advanced trainee based on the latest clinical guidelines, using a whiteboard animation software, with input from a multidisciplinary team. The development and description of the video is described following the Tidier Checklist in Supplement 2^15^.

The implementation strategy was the delivery of the intervention via a tablet with an internet connection, provided by the research assistant. The video was embedded in Redcap^18^ surveys (accessed via a weblink) and hosted in Vimeo account^19^.

### Recruitment and data collection

Eligible participants were identified through discussion with their care team and approached by the patient’s bedside. A research assistant performed the consent process and stayed with the patient while they watched the video. Patients were encouraged to provide feedback on the video during and after visualisation; the research assistant took notes of patients’ comments and feedback. At baseline, sociodemographic and MI knowledge data were collected. MI knowledge was assessed immediately post-intervention and at one month follow up (the latter being optional, for participants who opted to provide an email address for this purpose), as well as data on acceptability of the video. Sociodemographic data, MI knowledge, and acceptability data were collected via self-reported electronic questionnaires (hosted on RedCap ^18^ platform).

### Study Outcomes

The primary outcome was prospectively defined as the average number of correct responses in the MI knowledge questionnaire immediately after the intervention, compared to baseline. The MI knowledge questionnaire, developed by one of the investigators (AT), is a 10-item multiple choice (5 options with one correct answer per question) questionnaire that assesses general MI knowledge with questions on the causes of MI and post-MI medications (with the total number of correct answers ranging from 0, indicating no correct answers, to 10, meaning all answers to the 10 questions were correct) (Supplement 5).

Secondary outcomes included the proportion of participants who improved their total number of correct answers from baseline to post-intervention, the proportion meeting the knowledge target (defined as 7 or more correct responses out of 10), the proportion of correct responses for each of the ten individual questions post-intervention compared to baseline and the average number of correct responses in the MI knowledge questionnaire at one month.

To assess implementation, we evaluated the acceptability of the intervention and fidelity of the intervention and implementation strategy (i.e., mode of delivery of the intervention via a tablet). Acceptability of the intervention was assessed through thematic analysis of participants’ qualitative feedback (described in Data Analysis, below) and via a questionnaire asking participants to rate three different statements on a 5-point scale from strongly disagree to strongly agree (“The information delivered in the video was easy enough to understand”; “I found the video engaging”; and “I found the information useful”) and asking whether they would be interested in receiving similar videos in the future (Yes/No).

Fidelity of the intervention was assessed by the investigators as low, medium or high based on whether the core components of the intervention (i.e., video animations and educational content conveyed through the audio and subtitles) were delivered as intended across the four different language versions. For this assessment, perspectives from the translators involved in the study were sought regarding the extent to which the translated content matched the English version. Fidelity of the implementation strategy was assessed by the investigators as low, medium or high, based on research assistant notes of any deviations from the mode of delivery (tablet).

### Data Analysis

Data analysis followed a statistical analysis plan developed a priori (<u>Open Science Framework</u>). A sample size of 119 participants was estimated to provide 90% power to detect a moderate effect size (d=0.3) in knowledge difference pre-post, considering a 2-sided type 1 error of 0.05.

Descriptive data was expressed as proportions and means with standard deviation. The primary outcome was analysed using a paired t-test, measuring the mean difference in patient knowledge, before and after the intervention, reported with 95% confidence intervals. McNemar test was used to assess changes in the proportion of participants who met knowledge targets before and immediately post-intervention, proportion of correct responses for each of the ten individual questions post-intervention compared to baseline. A paired t-test was used to measure the mean difference in patient knowledge, before and one-month post-intervention. To test the homogeneity of the treatment effect across subgroups, we used a two-sample t-test to assess the change in average number of correct responses by age (≤ 65 years old and > 65 years old), sex (male and female), language (English and non-English) and education (prior to secondary education completion and post-secondary education). We report the proportion of participants who improved their total number of correct answers from baseline to post-intervention and implementation measures of acceptability descriptively immediately post-intervention and at one month follow-up.

Analyses were conducted using R statistical software version 4.4.0 (R Project for Statistical Computing).^20^ P value was 2-sided and statistical significance was set at p=0.05. Data were analysed from October 2024 to November 2024.

Two researchers performed thematic analysis on the notes taken of participants’ feedback.^21^ We were interested in understanding the participants’ perspectives on the content of the video in the different languages and their suggestions for improving the videos. Themes were reviewed and discussed with two other investigators to clarify, explore and refine interpretations.

## Results

Between December 2023 to October 2024, a total of 150 patients were assessed for eligibility, 130 were approached (20 were deemed by a clinician as not medically fit to be approached for the study), and 129 were enrolled (Figure 1). Of the 129 participants enrolled, 128 completed follow-up immediately post-intervention (one lost interest). Of the 89 participants who provided their email address for the one-month survey, 18 responded (Figure 1). The baseline characteristics of participants are presented in Table 1. The mean participant age was 59.4 years (SD 12.6) and 20.2% (26/129) were female. In our sample, 24.8% (32/129) were South Asian and 72.9% (94/129) completed secondary education or above. Majority of our participants listed English as their preferred language (74.4%; 96/129), followed by Hindi (13.2%; 17/129), Arabic (7.8%; 10/129) and Chinese (simplified) (4.7%; 6/129).

**Figure 1.**
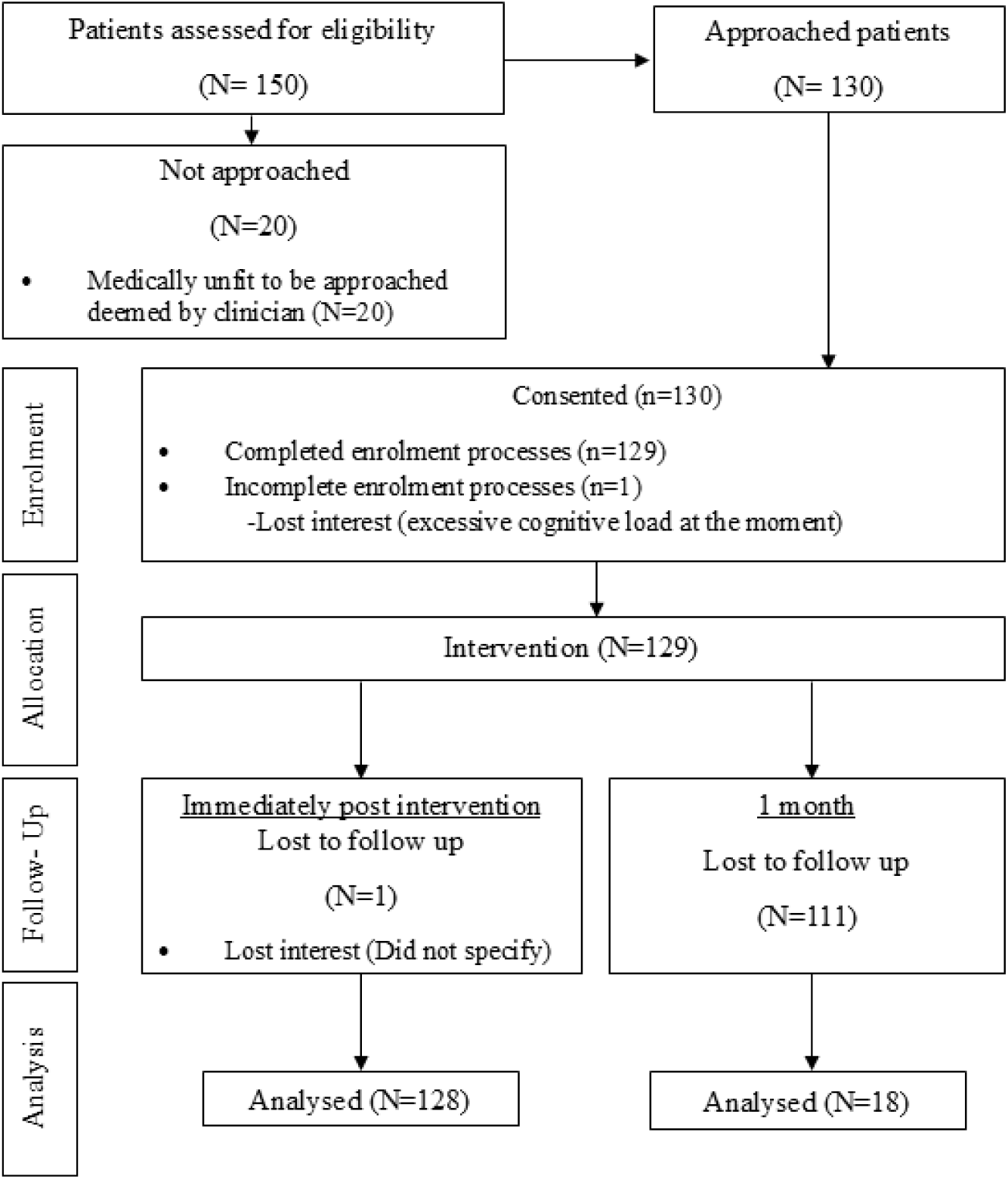
Consort Diagram

**Table 1.**
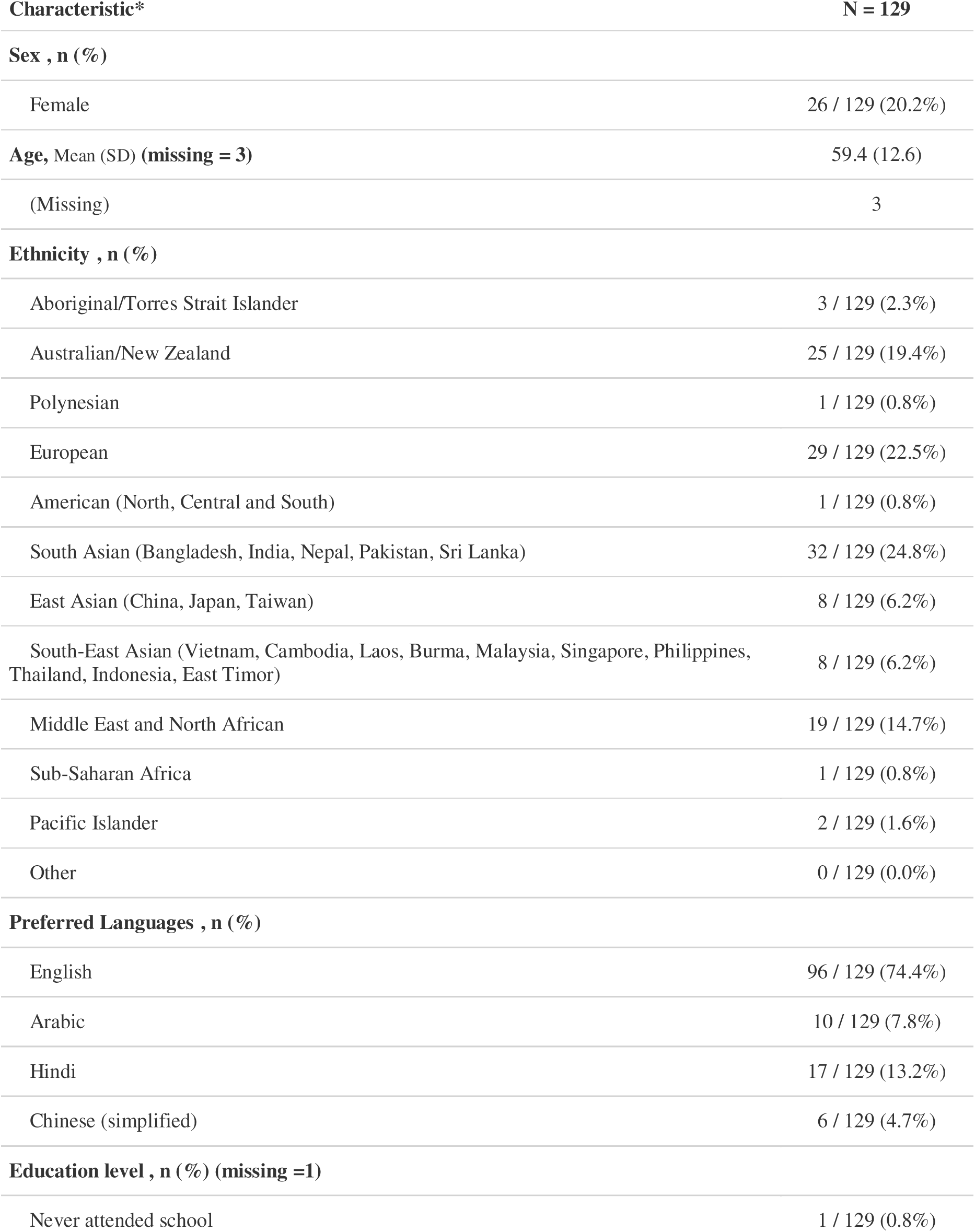
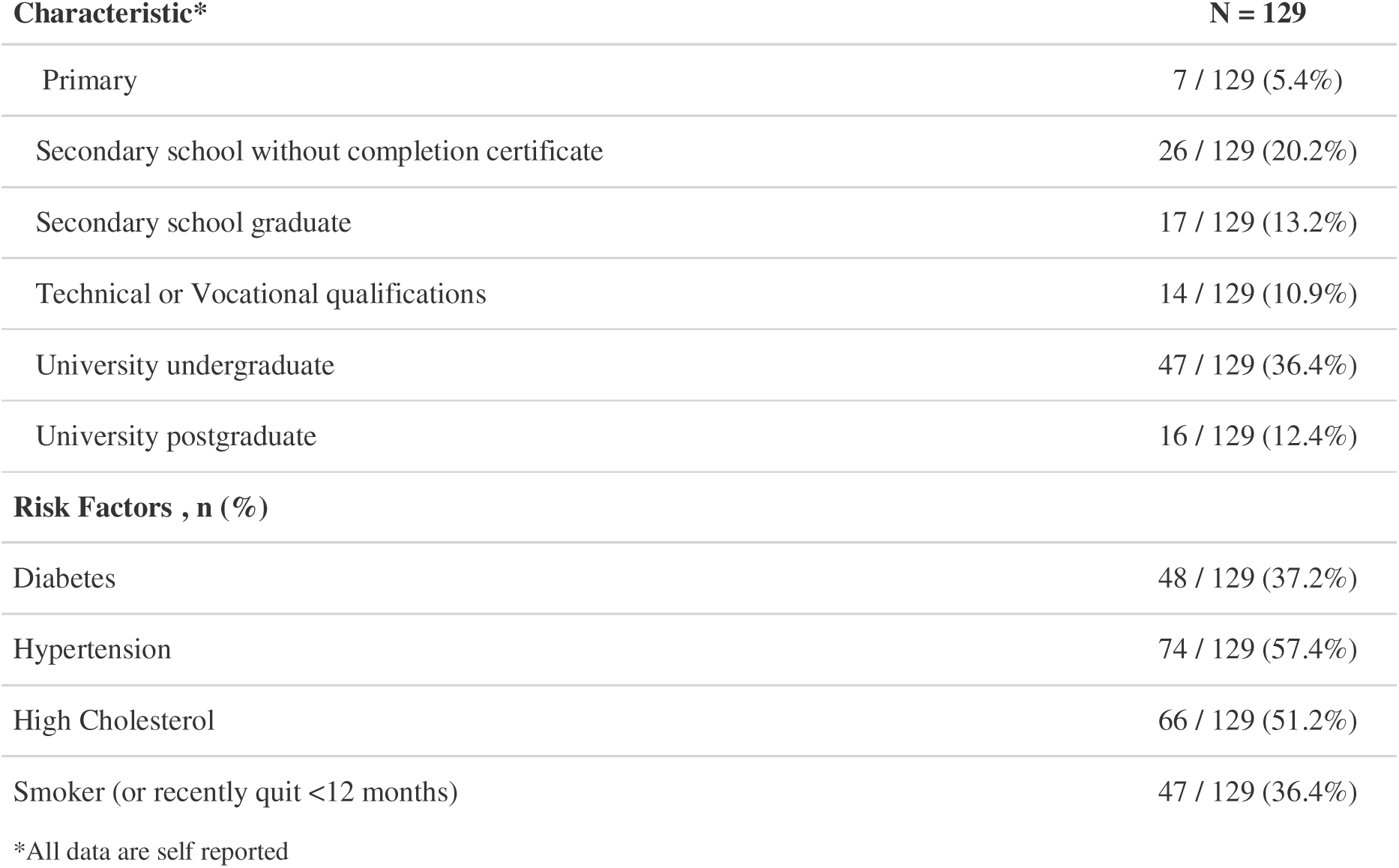
Baseline Characteristics.

The primary outcome analysis showed that the average number of correct responses increased from 5.4 (SD 2.7) at baseline to 7.2 (SD 2.5) immediately post-intervention (mean difference = 1.9; 95% CI 1.6, 2.2, p<0.001) (Table 2). Subgroup analysis by age, sex, language and education did not show significant differences in the average number of correct responses from baseline to immediate post-intervention (Supplement 6).

**Table 2.** Primary outcome analysis comparing the average number of correct responses baseline vs post-intervention.

|  | Mean (SD)<br>number of<br>correct<br>responses at<br>baseline<br>N=129 | Mean (SD) number<br>of correct responses<br>at immediately post<br>intervention<br>N=128 | Difference<br>N=128<br>[95% CI] | p-value |
| --- | --- | --- | --- | --- |
| **Number of**<br>**correct**<br>**responses\*** | 5.4<br>(2.7) | 7.2<br>(2.5) | 1.9<br>(1.9)<br>[1.6, 2.2] | < 0.001 |
| *Correct responses are out of 10 |  |  |  |  |

Overall, 72.7% (93/128) of the participants improved their total number of correct responses from baseline to immediately post-intervention (Supplement 7). The proportion of participants who met the MI knowledge target post-intervention, increased from 47/129 (36.4%) to 93/128 (72.7%) (p<0.001) (Figure 2, Table 3). For each of the individual questions, the proportion of correct responses increased from baseline to post-intervention, except for one question (Supplement 8-17).

**Figure 2.**
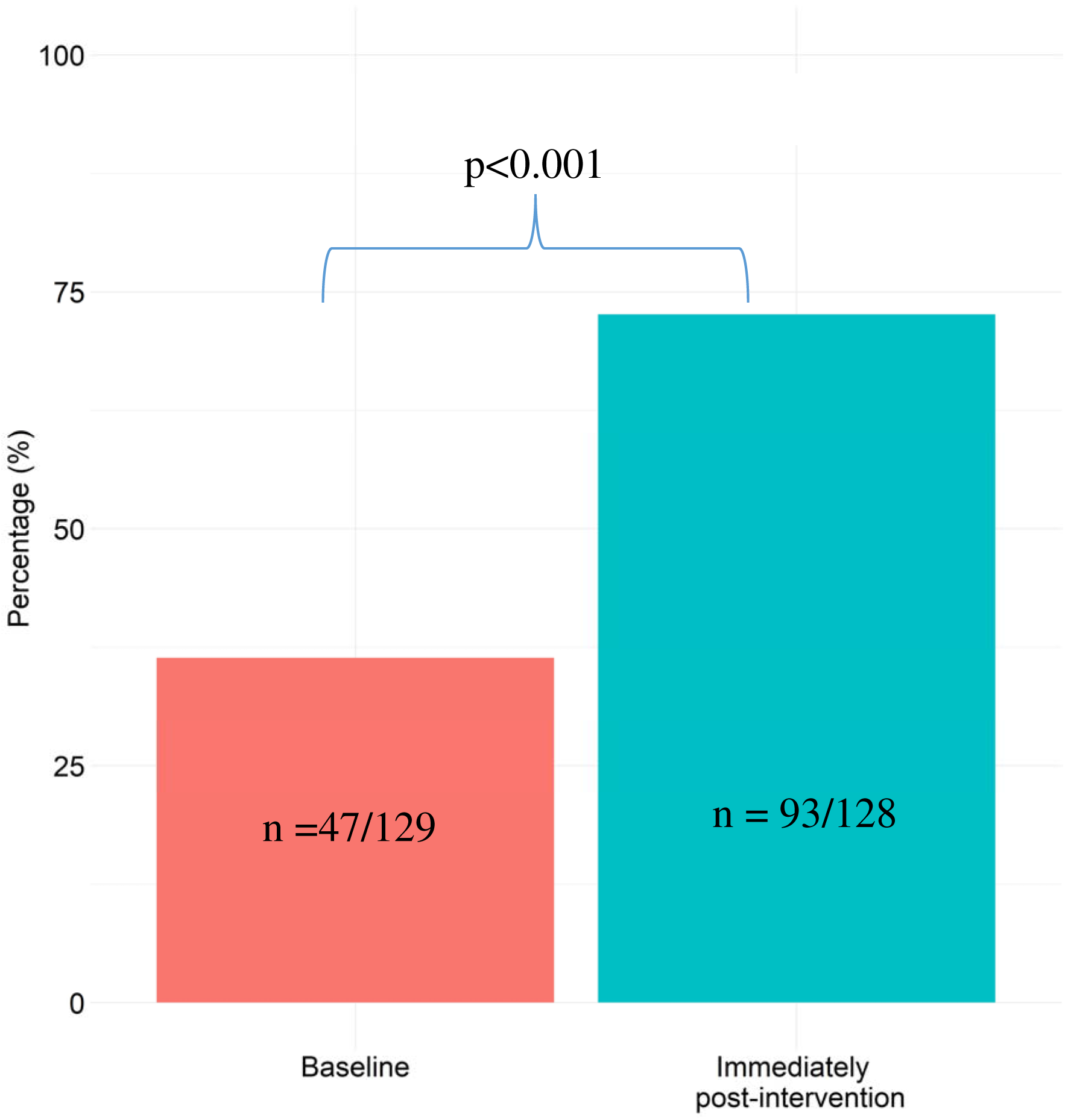
Meeting knowledge target (7/10 correct responses) baseline vs post-intervention descriptive analysis

**Table 3.** Proportion of participants who met general knowledge (7/10 correct responses) target baseline vs post-intervention.

|  | **Baseline, n / N (%)**<br>**N=129<sup>I</sup>** | **Immediately post**<br>**intervention, n / N**<br>**(%)**<br>**N=128<sup>I</sup>** | **p-value** |
| --- | --- | --- | --- |
| **Proportion of**<br>**participants who met**<br>**knowledge target (7/10-**<br>**number of correct**<br>**responses)** | 47/129 (36.4%) | 93 / 128 (72.7%) | < 0.001 |

At 1-month post-intervention, there were 18 completed responses. The average number of correct responses increased from 5.4 (SD 2.7) at baseline to 8.8 (SD 1.6) at one month (mean difference = 2.5; 95% CI 1.7, 3.3, p<0.001) (Supplement 18).

Regarding the acceptability of the intervention, most participants reported they agreed or strongly agreed the information delivered in the video was easy to understand (83.6%; 107/128), engaging (74.2%; 95/128), and useful (87.5%; 112/128), and 84.4% (108/128) reported they would like to receive similar videos in future (Supplement 19).

Nine participants (5 Hindi speaking, 2 Mandarin speaking, 2 Arabic speaking) provided qualitative feedback on the videos. Thematic analysis revealed two main themes: Complexity of the content and medical jargon; and Preference for conversational language and dialects. Regarding the complexity of the content and medical jargon, participants indicated they had difficulty understanding some terms in the videos, expressing a preference for lay language. Examples of quotes from participants grouped in this theme include:

> “The video will be too difficult to understand [for people not] familiar with the terminology” (Study ID 125; age 70-75; preferred language: Hindi; educational level: university undergraduate).

> “High order Hindi was used-needs to have less jargon” (Study ID 30; age 66-70; preferred language: Hindi; educational level: university postgraduate).

Regarding the preference for conversational language and dialects, participants with Hindi as their preferred language reported a macaronic-hybrid language was commonly used to communicate and would be preferred for the video. Participants with Arabic as their preferred language mentioned that they would prefer the video to be in their specific dialect. Examples of quotes are presented below:

> “Mix of Hindi and English would have been better - conversational Hindi” (Study ID 30; 66-70 years old; preferred language: Hindi; educational level: university postgraduate).

> “Dialect is like Egyptian … parents would understand Nahw … (I) am bilingual – so can’t read or write” (Study ID 108; 40-45 years old; preferred language: Arabic; educational level: university undergraduate).

Fidelity of the intervention was assessed as high, given the core components of the intervention (i.e. animations and educational content conveyed through the audio and subtitles) were delivered as intended. The video animations were the same across the four translated versions of the educational video, only the timing between different animations changed to accommodate the audio. The educational content conveyed through the audio and subtitles was also delivered as intended, despite minor nuances in language between the different translations. Fidelity of the implementation strategy was assessed as high because the delivery of the intervention was consistent for all patients (i.e. all patients watched the video on a tablet).

## Discussion

In this single-arm experimental trial of 129 inpatients admitted for an MI, a multilingual educational video significantly improved patient knowledge in the short term. Subgroup analyses did not show differences between participants whose preferred language was English versus other languages or between those with or without secondary education. The video was deemed acceptable, with most participants finding the videos easy to understand, engaging and useful. Fidelity of the intervention was high across the different language versions of the video, and the fidelity of the implementation strategy was also high, with consistent delivery of the video via a tablet. Our study demonstrates it is feasible to deliver videos in different languages (English, Arabic, Hindi and Mandarin) to patients in a multicultural context at a tertiary hospital.

Results from our study suggest that a single video intervention can improve patient MI knowledge across different language groups in the inpatient setting. Our results are consistent with other studies evaluating inpatient video interventions for patients post-MI^12,13^, although ours was the only one delivered in more than one language. One of these studies, an RCT of 68 inpatients, demonstrated a significant within-group improvement in knowledge at 3 months in the group receiving the 15-minute video, but no difference when compared to standard education^12^. The other study (quasi-experimental; 2-arm; n=25) showed a significant improvement in knowledge in the group receiving the 7-minute video, compared to standard education.^13^ Multilingual educational videos have been evaluated in other health domains, such as in the context of intravenous contrast administration consent (RCT; n=112) ^22^, and human papillomavirus vaccine education (RCT; n=708)^23^, both showing significant improvements in knowledge in the intervention group when compared to control, with no differences between intervention participants receiving the English versus non-English video. It is unclear how different health literacy strategies applied to multilingual videos can influence knowledge in different populations, which should be explored in future research^24^.

The mean post-intervention score was 7/10, even after participants had just viewed the video, highlighting the persistent challenges of meeting patients’ educational and health literacy needs. These findings were echoed in our feedback discussions and may reflect broader barriers to comprehending health resources, such as high readability levels (i.e., above the recommended Grade 8 level) and the use of medical jargon^25^. Reducing readability levels prior to translation into non-English languages is important, as content complexity may be compounded with translation ^25–28^. In addition, the literal translation of certain terms may not accurately convey their intended meaning, and the use of culturally equivalent terms may be preferable^25–29^. Hence, cultural adaptation is key to adequately considering the cultural aspects that may not be captured through linguistic translation alone, such as cultural equivalence, cultural appropriateness, similarity of interpretability, and item relevance ^25,30–32^.

While knowledge scores improved post-intervention and the video in our study was well-accepted, further gains may be achieved with the involvement of community members in the co-design and cultural adaptation of intervention content^25,32,33^. Notwithstanding the importance of addressing language barriers in improving health outcomes and quality of care^34,35^, future studies aiming to develop equitable health education interventions should also consider the nuances of social context and how language intersects with race, migration status, religion, socioeconomic position, and other social determinants of health^36–38^.

### Strengths and limitations

Strengths of this study include delivering the intervention to a diverse multicultural population (88% non-Caucasian) in an inpatient setting. We translated the intervention into four different languages and adapted medical jargon in the translations. However, there were challenges in translating medical language, and the readability of our English-language content was high, at Grade 10, instead of the desired Grade 8.

This study should be interpreted in the context of some limitations. Only 34/129 participants received the intervention in a language other than English, and the qualitative feedback from these patients had to be provided in English or Mandarin to the bilingual research assistant, so important feedback may have been missed. Patient knowledge may have been influenced by recall in the immediate post-intervention, and the one-month results should be interpreted with caution due to low response rates. The MI knowledge questionnaire was not validated; it was created by clinicians in the study, given the absence of validated patient questionnaires to assess myocardial infarction knowledge. Finally, we did not assess health literacy or language proficiency, which could have aided in the interpretation of the results.

### Implications – for research, practice and policy

Our study has the potential to reduce language barriers in the delivery of inpatient education for patients post-MI. We demonstrated that it is possible to improve knowledge in the short term and implement education videos within a 5-minute timeframe for hospitalised MI patients in a multicultural context, offering a scalable strategy that may ease demands on healthcare staff. Patients report a considerable treatment burden in the first year following an MI, particularly those with lower health literacy^39^, which may contribute to suboptimal medication adherence^40^. Providing education prior to discharge may help alleviate this burden and serve as a primer for adherence to secondary prevention care. In the future, large language models and other generative artificial intelligence (AI) tools could complement video-based education by providing interactive, personalised support^41^ in a patient’s preferred language^42–44^ and at an appropriate readability level^45^. These tools could enhance health literacy and support equitable care for patients from diverse linguistic backgrounds.

## Conclusions

Multilingual videos delivered during inpatient hospital admission for acute-MI may improve patient knowledge in the short term. Our study demonstrates it is feasible to deliver videos in different languages (English, Arabic, Hindi and Mandarin) to patients in a multicultural context at a tertiary hospital. Further research is needed to evaluate the long-term impact of this intervention on MI knowledge and secondary prevention behaviours across different settings.

## Data Availability

The datasets generated and/or analyzed during this study can be made available from the corresponding author on reasonable request.

## Acknowledgements

We acknowledge research officer, Wendy Liu’s contribution for her role in speaking the Mandarin voiceover for one of the videos and medical officer, Dr Rukmini Kulkarni’s contribution for her role in speaking the Hindi voiceover. We acknowledge the nursing staff and doctors from Westmead Cardiology Unit for facilitating recruitment efforts and the biostatistical team at Westmead Applied Research Centre (Haeri Min and Dr Desi Quintans) for validating our results.

## Abbreviations

CVD: Cardiovascular disease
MI: Myocardial infarction

